# Extrahepatic multimorbidity, disease clusters and overall and cause-specific mortality in people with steatotic liver diseases: a prospective analysis of UK Biobank

**DOI:** 10.1101/2025.03.06.25323330

**Authors:** Qi Feng, Chioma Izzi-Engbaeya, Pinelopi Manousou, Mark Woodward

## Abstract

**Background:** Steatotic liver disease (SLD) is a prevalent chronic liver disease linked to increased risks of various liver and extrahepatic diseases. However, the clustering of extrahepatic conditions and their impact on mortality in individuals with SLD remain poorly understood.

**Methods:** We used UK Biobank data to identify disease clusters among individuals with SLD and multimorbidity (having >= 2 extrahepatic diseases) using latent class analysis, conducted separately in males and females. Multivariable Cox models were used to assess associations between multimorbidity, derived disease clusters and all-cause mortality and cause-specific mortality of cardiovascular diseases, extrahepatic cancers, hepatocellular carcinoma and liver related diseases, with individuals without multimorbidity as reference group.

**Results:** Among 36002 SLD individuals with multimorbidity, we identified five disease clusters in both sexes: respiratory, mental health, cancer/osteoarthritis, and cardiovascular clusters. Males had separate heart and stroke clusters, whereas females had a combined heart/stroke cluster and a unique thyroid cluster. During a median follow-up of 13.8 years, cardiovascular disease was the leading cause of death in cardiovascular clusters, whereas extrahepatic cancers were the most common cause of death in other clusters. Multimorbidity was associated with increased mortality by 100% (HR (95%CI): 2.00 (1.93, 2.08)), as well as mortality of cardiovascular diseases, extrahepatic cancers, and liver-related diseases. Among all disease clusters, cardiovascular clusters exhibited the highest mortality risk: 2.36 (2.16, 2.58) for the stroke cluster, 2.63 (2.48, 2.78) for the heart cluster in males, and 2.90 (2.64, 3.20) for heart/stroke cluster in females.

**Conclusions:** Multimorbidity clusters in SLD exhibit sex differences, with cardiovascular-related clusters showing the highest mortality. These findings highlight the need for tailored prevention and management strategies in SLD populations.

## Introduction

Steatotic liver disease (SLD) is the most prevalent chronic liver disease, affecting every one in three adults worldwide [1]. SLD can progress to liver fibrosis, cirrhosis and hepatocellular carcinoma (HCC) and end-stage liver disease [2,3]. SLD is widely considered as a hepatic manifestation of broad metabolic dysfunction, with close links to cardiometabolic risk factors, such as obesity, hypertension, diabetes and dyslipidaemia [4]. Beyond liver-related conditions, it is also associated with increased risk of cardiovascular diseases (CVD), extrahepatic cancers and chronic kidney disease [5–7].

Multimorbidity (MM), the coexistence of two or more long-term conditions (LTCs), is increasingly recognized as a major public health challenge, particularly in aging populations [8,9]. People with SLD frequently exhibit multimorbidity, largely driven by cardiometabolic and cancer comorbidities. However, beyond these well-established associations, the broader multimorbidity patterns in individuals with SLD remain poorly characterized. Understanding these disease clusters is crucial for improving risk stratification, understanding mechanisms, optimising clinical management and healthcare services, thus improving individual long-term outcomes [10].

Previous studies have used various clustering techniques to derive disease clusters in general population, often identifying cardiometabolic, respiratory, and mental health clusters [11,12]. However, few studies have specifically examined extrahepatic multimorbidity clusters in individuals with SLD. Given the high burden of liver-related and extrahepatic complications in this population, it is essential to determine whether distinct disease clusters exist and how they may impact mortality outcomes. Additionally, sex differences in disease clustering remain underexplored [8].

This study was aimed to identify multimorbidity clusters among individuals with SLD in the UK Biobank, and to examine the associations between these clusters and all-cause and cause-specific mortality, considering sex differences in disease clustering and outcomes.

## Methods

### Data and participants

We used data from UK Biobank, a prospective cohort of half million individuals aged between 40 and 70 years old recruited between 2006 and 2010. Baseline assessment collected data on socioeconomic status, lifestyle factors, health status, occupational and environmental exposures, and anthropometric measures. Biological samples of blood, urine and saliva were collected. All participants were followed up via linkage to national death registries and hospital records.

We included people with SLD, defined using a fatty liver index (FLI) > 60 as an indicator of liver steatosis. FLI is a biomarker-based score for hepatic steatosis levels, calculated with body mass index (BMI), waist circumference, triglycerides (TG) and gamma-glutamyl transferase (GGT) [13]. The cutoff value of 60% has been validated and used previously to define liver steatosis [14,15]. SLD subtypes were classified into metabolic dysfunction-associated steatotic liver disease (MASLD), metabolic dysfunction and alcohol related liver disease (MetALD), alcohol related liver disease (ALD) and others, based on alcohol consumption and the presence of cardiometabolic risk factors (CMRF) [3,16]. CMRFs included obesity, hypertension, diabetes, high TG and low high-density lipoprotein (HDL) cholesterol, and we measured these CMRFs as defined in Rinella et al. [16]. We excluded people who withdrew from the cohort, who were pregnant at baseline, and people who had missing data in calculating FLI.

### Multimorbidity and disease clusters

We defined multimorbidity (MM) as presence of at least two LTCs of 47 diseases from a prespecified list. This list of disease was based on a three-round Delphi study of healthcare professionals and public members [17]. The criteria for including these LTCs included their impacts on mortality, quality of life, frailty, physical disability, mental health and treatment burden. Among the conditions identified in this Delphi study, we combined solid organ cancers, metastatic cancers, melanoma, and treated cancer requiring surveillance, into one broad solid organ cancer category, and removed post-acute Covid-19, chronic Lyme disease, and two liver conditions (hepatocellular carcinoma (HCC) and chronic liver disease). Since SLD is our index disease, we excluded SLD and its associated CMRFs from the list. After these modifications, this study considered 47 LTCs for multimorbidity (supplementary methods). These LTCs covered extrahepatic cancers, cardiovascular, metabolic, endocrinological, respiratory, digestive, renal, mental/behavioural and congenital conditions. Diagnosis of these conditions in each individual was confirmed in self-report medical conditions during nurse-led verbal interview at baseline and hospital records on and before the baseline assessment.

We used latent class analysis [18] to determine disease clusters in individuals with multimorbidity (>= 2 LTCs), separately in males and females, as previous evidence has suggested sex differences in disease clusters [11]. Latent class analysis allocates each individual with multimorbidity to a single non-overlapping cluster, and allows each condition to contribute to various clusters by varying probabilities. We compared the performance statistics for multiple latent class analysis model of between 2 to 12 clusters, using Bayesian Information Criteria (BIC), sample-size adjusted BIC and entropy statistics. The optimal number of clusters was decided using a combination of model performance statistics, clinical judgement, and capping the smallest cluster to great than 5% of the sample [11,12].

The derived clusters were characterised by the 3 LTCs with the highest probabilities great than 5% of contributing to that cluster, excluding condition for which their observed prevalence was equal to or less than that of the expected prevalence of the total sample [11].

### Outcome and covariates

The outcomes were all-cause mortality and mortality of HCC, liver related diseases, CVD and extrahepatic cancers. Cause of death was confirmed in death registry records. participants were censored at the date of death or the last day of follow-up (30 November 2022), whichever occurred earlier. We used the following ICD10 codes for specific causes of death: HCC (C22.0), liver-related diseases (K70-K77, C22), CVD (I00-I99), and extrahepatic cancer (C00-C99 excluding C22). Liver-related diseases included chronic liver disease (K70-K77) and liver cancer (C22).

A participant’s region was determined by the location of the assessment centre they attended. Ethnicity was classified into White, Asian, Black, and mixed/others. The Townsend Deprivation Index is a postcode-derived measure for socioeconomic status. Educational attainment was categorised as below secondary, lower secondary, upper secondary, vocational training, and higher education. Lifestyle factors included self-reported current smoking (current, previous and never smoker), alcohol consumption, and physical activity level. Alcohol consumption was assessed via self-reported weekly or monthly intake of various alcoholic drinks; the consumption was summed up to derive average daily alcohol consumption (g/d) [19]. Physical activity level was measured with the International Physical Activity Questionnaire; individuals were categorized into low, moderate and high levels, based on the frequency, duration and intensity of their physical activities. Systolic and diastolic blood pressure were measured twice and the averages of the two readings were used in analyses. Blood biochemistry markers were measured at a central laboratory, including TG, HDL-cholesterol, HbA1c, ALT, AST and GGT. For all the categorical covariates, answers of “unknown”, “do not know”, “prefer not to say” were combined into one “unknown” category.

### Statistical analysis

The baseline characteristics of males and females with SLD were summarised and compared between those with multimorbidity and those without, as well as between the derived disease clusters. To identify risk factors for each cluster, we fitted multinominal logistic regression, with age, ethnicity, physical activity, smoking, alcohol intake (per 10 g/day), and CMRFs.

Cox proportional hazard models were fitted to assess the associations between multimorbidity, disease clusters and mortality outcomes, expressed as hazard ratio (HR) and 95% confidence interval (CI), using individuals without multimorbidity as the reference group. The models were stratified by region and age groups, and adjusted for age, ethnicity, Townsend Deprivation Index (in fifth), education, smoking, physical activity, and alcohol cunsumption. The proportional hazard assumption was examined by scaled Schoenfeld residuals, and no evidence was observed for its violation. For sensitivity analyses, we (1) further adjusted for the five CMRFs, and (2) removed the first two years of follow-up to correct for reverse causation.

All analyses were done in R.

## Results

After exclusion, we included 178335 individuals with SLD in analysis (36.3% females, mean age 57.3 years, 73.5% MASLD, 19.0% MetALD, 6.4% ALD) (figure 1). About one in five (20.2%) of SLD participants had MM, which was higher in females than in males (24.4% vs. 17.8%). Overall, compared to people without MM, people with MM were more likely to be older, socioeconomically deprived, less educated, smokers and drinking less. They were also more likely to have diabetes and low HDL-cholesterol, but less likely to have hypertension and high TG. (table 1)

**Figure 1:**
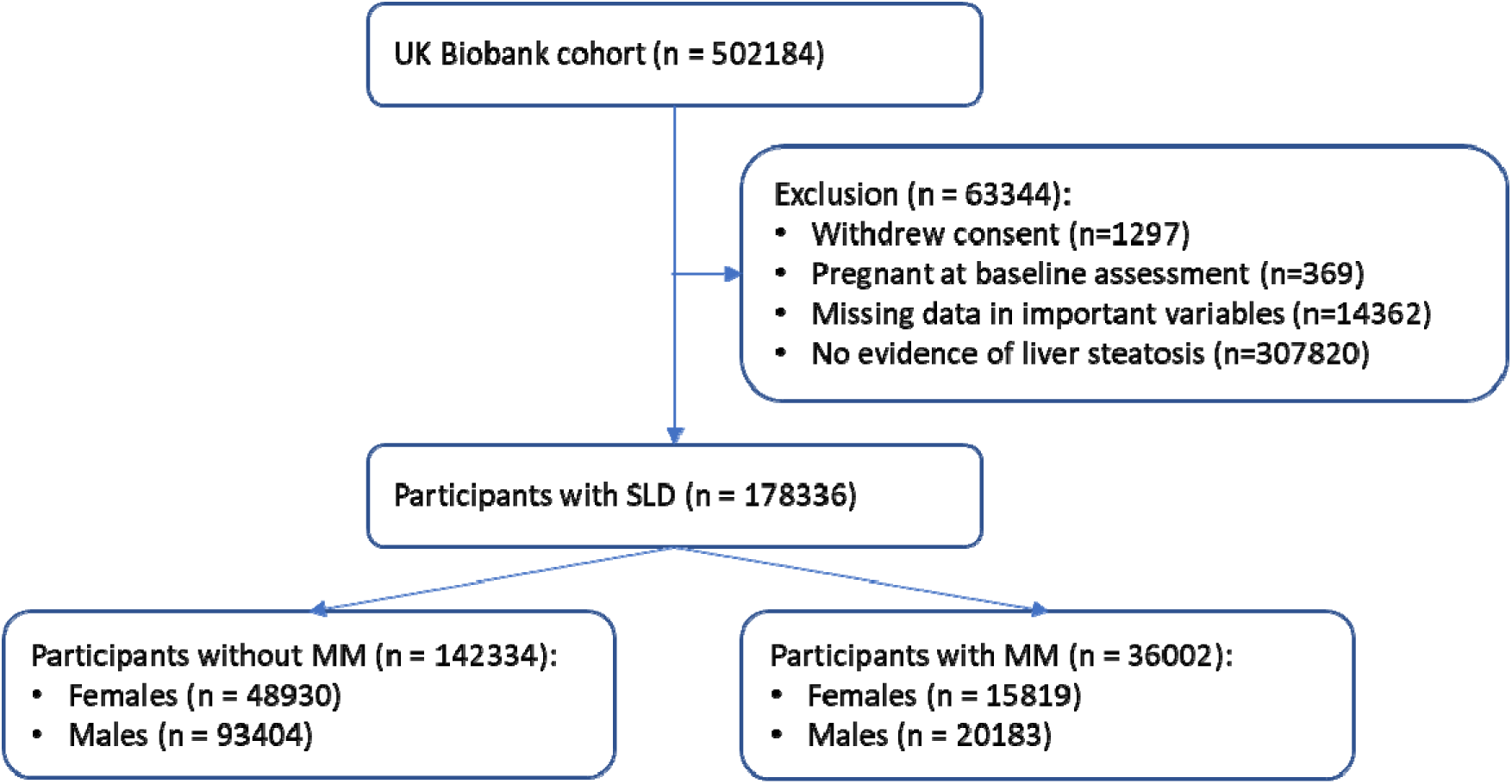
Flowchart of patient selection. SLD: steatotic liver disease. MM: multimorbidity.

**Table 1:** Baseline characteristics in participants with steatotic liver disease stratified by sex and multimorbidity status.

|  | Males |  |  | Females |  |  |
| --- | --- | --- | --- | --- | --- | --- |
| | $\geq 2$ LTCs | $< 2$ LTC | | $\geq 2$ LTCs | $< 2$ LTC | Overall |
|  | n = 20183 | n = 93404 |  | n = 15819 | n = 48930 | n = 178336 |
| **Age, years** | 59.4 (7.5) | 56.5 (8.0) |  | 58.6 (7.4) | 57.4 (7.5) | 57.3 (7.8) |
| **Townsend deprivation index** |  |  |  |  |  |  |
| 1st fifth (least deprived) | 3188 (15.8%) | 18637 (20.0%) |  | 2188 (13.8%) | 8449 (17.3%) | 32462 (18.2%) |
| 5th fifth (most deprived) | 5728 (28.4%) | 18804 (20.1%) |  | 4853 (30.7%) | 11485 (23.5%) | 40870 (22.9%) |
| **Education**, higher education | 4727 (23.4%) | 27880 (29.8%) |  | 3157 (20.0%) | 11582 (23.7%) | 47346 (26.5%) |
| Ethnicity, White | 19263 (95.4%) | 88343 (94.6%) |  | 14853 (93.9%) | 45724 (93.4%) | 168183 (94.3%) |
| **Smoking**, never | 7254 (35.9%) | 42439 (45.4%) |  | 7983 (50.5%) | 27847 (56.9%) | 85523 (48%) |
| Alcohol drinking, g/day | 22.9 (27.8) | 25.6 (26.0) |  | 7.3 (12.9) | 9.0 (13.1) | 19.1 (23.8) |
| Physical activity, high | 5142 (25.5%) | 29056 (31.1%) |  | 3042 (19.2%) | 10595 (21.7%) | 47835 (26.8%) |
| BMI, kg/m<sup>2</sup> | 31.1 (4.4) | 30.3 (3.7) |  | 33.9 (5.3) | 33.2 (4.7) | 31.5 (4.5) |
| FIB4 score | 1.6 (6.8) | 1.4 (1.0) |  | 1.3 (3.5) | 1.2 (1.3) | 1.3 (2.7) |
| **Waist circumference, cm** | 106.4 (10.8) | 103.4 (9.5) |  | 101.7 (10.8) | 99.6 (9.7) | 102.6 (10.0) |
| Systolic blood pressure, mmHg | 141.3 (17.6) | 143.9 (17.0) |  | 139.6 (18.0) | 141.5 (18.3) | 142.6 (17.6) |
| Diastolic blood pressure, mmHg | 83.9 (10.3) | 86.7 (9.7) |  | 83.6 (9.9) | 85.2 (9.5) | 85.7 (9.8) |
| ALT, log<sub>10</sub> U/L | 1.4 (0.2) | 1.5 (0.2) |  | 1.4 (0.2) | 1.4 (0.2) | 1.4 (0.2) |
| AST, log<sub>10</sub> U/L | 1.4 (0.2) | 1.4 (0.1) |  | 1.4 (0.2) | 1.4 (0.1) | 1.4 (0.2) |
| GGT, log<sub>10</sub> U/L | 1.7 (0.3) | 1.7 (0.3) |  | 1.6 (0.3) | 1.6 (0.3) | 1.6 (0.3) |
| Triglycerides, mmol/L | 2.1 [1.4] | 2.2 [1.4] |  | 2.0 [1.2] | 2.0 [1.2] | 2.1 [1.3] |
| HDL cholesterol, mmol/L | 1.1 (0.4) | 1.1 (0.4) |  | 1.3 (0.4) | 1.3 (0.4) | 1.2 (0.4) |
| HbA1c, mmol/mol | 38.5 (11.6) | 36.2 (9.3) |  | 38.1 (10.6) | 36.8 (9.3) | 36.8 (9.8) |
| Platelet, 10<sup>9</sup> cells/L | 237.7 (60.9) | 238.7 (54.9) |  | 277.1 (66.3) | 274.7 (61.4) | 251.8 (61.1) |
| Hypertension | 15450 (76.5%) | 77040 (82.5%) |  | 13038 (82.4%) | 39889 (81.5%) | 145417 (81.5%) |
| Obesity | 19788 (98.0%) | 91381 (97.8%) |  | 15788 (99.8%) | 48837 (99.8%) | 175794 (98.6%) |
| Diabetes | 7795 (38.6%) | 23050 (24.7%) |  | 6182 (39.1%) | 14735 (30.1%) | 51762 (29.0%) |
| High triglyceride | 13759 (68.2%) | 67561 (72.3%) |  | 12287 (77.7%) | 34685 (70.9%) | 128292 (71.9%) |
| Low HDL-cholesterol | 7266 (36.0%) | 27621 (29.6%) |  | 10506 (66.4%) | 27784 (56.8%) | 73177 (41.0%) |

LTC: long-term condition. BMI: body mass index. FIB4: fibrosis-4 score. ALT: alanine aminotransferase. AST: aspartate aminotransferase. GGT: gamma-glutamyl transferase. HDL: high-density lipoprotein. HbA1c: glycated hemoglobin.

### Disease clusters and their characteristics

Latent class analysis derived five distinct disease clusters for males and females, respectively, with some overlapping and some sex-specific patterns (supplementary figure 1, supplementary table 1). Table 2 summarises the characteristic diseases within each cluster. In both sexes, a respiratory cluster (characterized by asthma, COPD, and other chronic respiratory diseases), a mental health cluster (dominated by depression and anxiety, with substance use disorder additionally present in males), and a cancer/osteoarthritis cluster (including solid organ cancers, chronic respiratory diseases, and osteoarthritis) were observed. Among males, a heart cluster (characterized by ischemic heart disease, arrhythmia, and heart failure) and a stroke cluster (characterised by stroke and paralysis) were identified. In females, these two clusters merged into a single heart/stroke cluster, which included ischemic heart disease, arrhythmia, and stroke. Additionally, females had a unique thyroid cluster, consisting of thyroid disorders and connective tissue diseases.

**Table 2:**
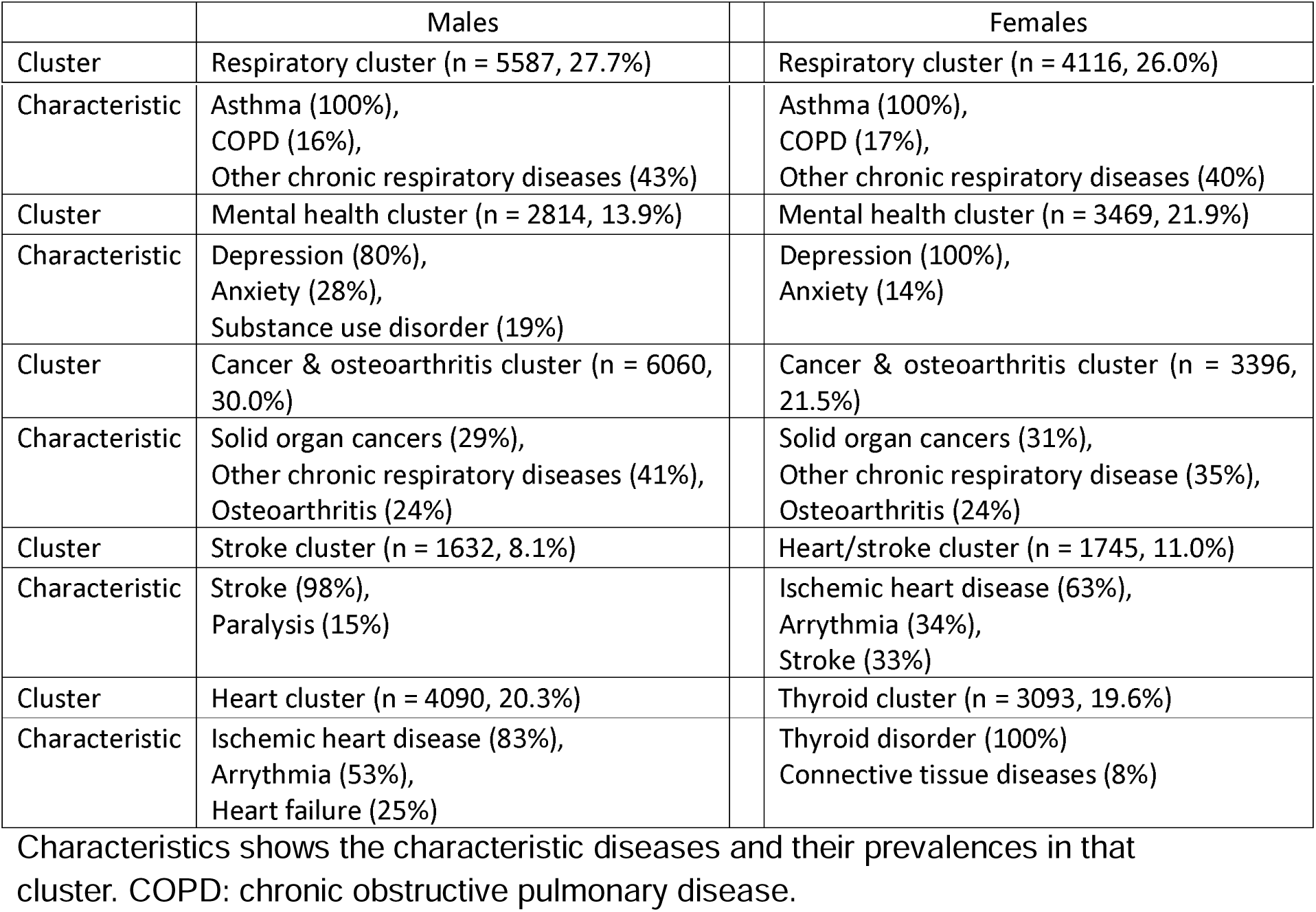
Latent class analysis derived disease clusters in males and females with steatotic liver disease.

|  | Males |  | Females |
| --- | --- | --- | --- |
| Cluster | Respiratory cluster (n = 5587, 27.7%) |  | Respiratory cluster (n = 4116, 26.0%) |

|  |  |  |
| --- | --- | --- |
| Characteristic | Asthma (100%),<br>COPD (16%),<br>Other chronic respiratory diseases (43%) | Asthma (100%),<br>COPD (17%),<br>Other chronic respiratory diseases (40%) |
| Cluster | Mental health cluster (n = 2814, 13.9%) | Mental health cluster (n = 3469, 21.9%) |
| Characteristic | Depression (80%),<br>Anxiety (28%),<br>Substance use disorder (19%) | Depression (100%),<br>Anxiety (14%) |
| Cluster | Cancer & osteoarthritis cluster (n = 6060, 30.0%) | Cancer & osteoarthritis cluster (n = 3396, 21.5%) |
| Characteristic | Solid organ cancers (29%),<br>Other chronic respiratory diseases (41%),<br>Osteoarthritis (24%) | Solid organ cancers (31%),<br>Other chronic respiratory disease (35%),<br>Osteoarthritis (24%) |
| Cluster | Stroke cluster (n = 1632, 8.1%) | Heart/stroke cluster (n = 1745, 11.0%) |
| Characteristic | Stroke (98%),<br>Paralysis (15%) | Ischemic heart disease (63%),<br>Arrhythmia (34%),<br>Stroke (33%) |
| Cluster | Heart cluster (n = 4090, 20.3%) | Thyroid cluster (n = 3093, 19.6%) |
| Characteristic | Ischemic heart disease (83%),<br>Arrhythmia (53%),<br>Heart failure (25%) | Thyroid disorder (100%)<br>Connective tissue diseases (8%) |

Characteristics shows the characteristic diseases and their prevalences in that cluster. COPD: chronic obstructive pulmonary disease.

In males, the most common disease cluster was the cancer/osteoarthritis cluster (30.0%), followed by respiratory cluster (27.7%), heart cluster (20.3%) and mental cluster (13.9%) while stroke cluster (8.1%) was the least common. In females, the most common cluster was respiratory cluster (26.0%), followed by mental cluster (21.9%), cancer/osteoarthritis cluster (21.5%) and thyroid cluster (19.6%), and the least common was heart/stroke cluster (11.0%) (table 2). These trends were generally consistent across all SLD subtypes (supplementary figure 2)

Supplementary table 2 and 3 present the baseline characteristics of males and females across the disease clusters. Supplementary table 4 and 5 show the results of multinominal logistic regression, with the respiratory cluster as the referent group. In both males and females, disease clustering was associated with age, socioeconomic status, lifestyle and CMRFs. In males, older age and socioeconomic deprivation were linked to higher odds of being in the cancer/osteoarthritic, stroke and heart clusters, while higher educational attainment was associated with lower odds of these clusters. Asian ethnicity was associated with lower prevalence of mental health cluster, while black ethnicity was associated with higher prevalence of cancer/osteoarthritis cluster. Physical activity was protective against the stroke cluster. Smoking and low HDL were consistently associated with higher odds of all disease cluster. Diabetes was found to increase the odds of heart and stroke clusters.

In females, older age was associated higher odds of cancer/osteoarthritis, heart/stroke and thyroid clusters, but lower odds of mental health cluster. Asian was associated with lower odds of mental health cluster, but higher odds of cancer/osteoarthritis, heart/stroke and thyroid clusters. Heart/stroke cluster was more prevalent in people with socioeconomic deprivation, smokers, those with lower education, diabetes and low HDL levels.

### Disease clusters and mortality

Among a median follow-up of 13.8 years, 14595 and 6171 deaths were captured in males and females, respectively. Compared to males without MM, males with MM had an excess mortality rate of 12.3/1000 person-years (20.0 vs. 7.7), yielding an HR estimate of 2.00 (95%CI 1.93, 2.08). Compared to females without MM, females with MM had an increased mortality by 80% (mortality rate 11.7 vs. 5.7/1000 person-years, HR 1.80 (1.71, 1.90)).

All disease clusters showed higher mortality than people without MM. In males, heart and stroke clusters were associated with the highest mortality, with HR of 2.63 (2.48, 2.78) and 2.36 (2.16, 2.58); respiratory, mental health and cancer/osteoarthritis clusters showed HR of 1.62 (1.51, 1.73), 1.84 (1.69, 2.00) and 1.85 (1.75, 1.96), respectively. In females, heart/stroke cluster was associated with the highest mortality, with HR of 2.90 (2.64, 3.20). Respiratory, mental health, cancer/osteoarthritis and thyroid clusters showed HR of 1.73 (1.58, 1.89), 1.57 (1.42, 1.74), 1.85 (1.69, 2.02) and 1.42 (1.28, 1.58), respectively (Figure 2, supplementary table 6). Sensitivity analyses by additional adjustment for CMRFs and removing the first two years of follow-up generated similar results (supplementary table 7).

**Figure 2:**
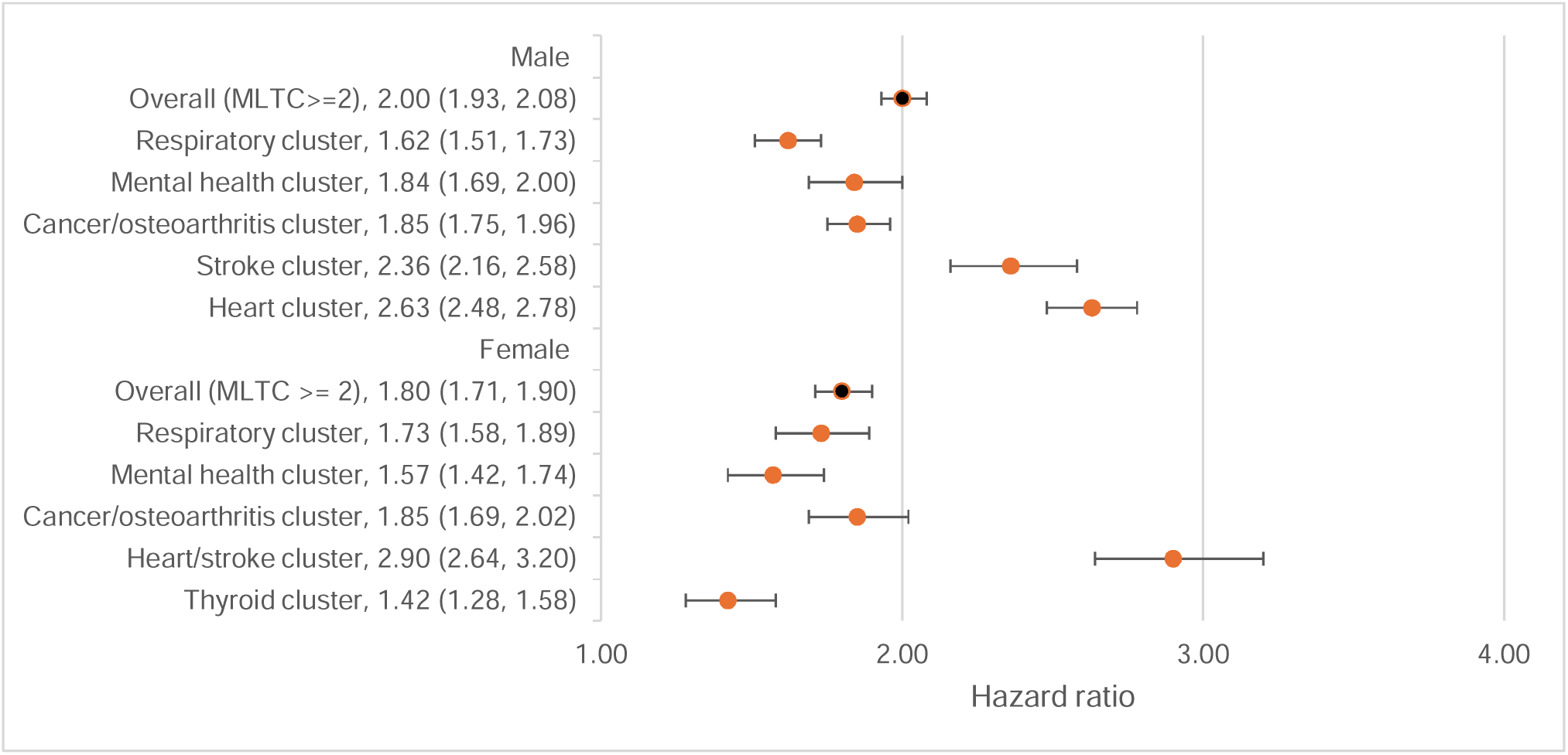
Associations between multimorbidity, disease clusters and all-cause mortality in males and females with steatotic liver disease. LTC: long-term conditions.

### Disease clusters and cause-specific mortality

Examining causes of death, in males, 31.9% and 31.8% deaths were attributed to extrahepatic cancers and CVD, while only 1.1% to HCC and 4.4% to liver related diseases. Similarly in females, 39.9%, 21.9%, 0.3% and 3.3% deaths were attributed to extrahepatic cancers, CVD, HCC and liver related diseases, respectively. Extrahepatic cancer and CVD were the major causes of death across all clusters, contributing 58.1% to 67.3% of all deaths in males and 59.1% to 66.7% in females. CVD was the biggest cause of death in the stroke and heart clusters in males and in the stroke/heart cluster in females, while extrahepatic cancers remained the biggest cause of death for all other clusters. (supplementary figure 3)

Compared to people without MM, having MM was associated with higher mortality of extrahepatic cancers by 45% (1.45 (1.37, 1.54)), CVD by 173% (2.73 (2.56, 2.92)), HCC by 67% (1.67 (1.20, 2.32)) and liver related diseases by 84% (1.84 (1.55, 2.18)) in males, and extrahepatic cancers by 40% (1.40 (1.30, 1.52)), CVD by 130% (2.30 (2.04, 2.58)), and liver related death by 46% (1.41 (1.06, 1.86)) in females. All five clusters were associated with mortality of extrahepatic cancers, with the highest risk in cancer clusters in males (1.77 (1.62, 1.92)) and females (1.74 (1.53, 1.98)). All five clusters were associated with mortality of CVD, with the highest risk in stroke and heart clusters in males (2.24 (2.86, 3.90) and 5.28 (4.83, 5.77)) and stroke/heart cluster (5.66 (4.75, 6.75)) in females. The disease clusters also showed general trend of positive associations with mortality of HCC and liver related diseases, although non-significantly probably due to small number of events in these clusters. (table 3)

**Table 3:** Associations between the disease clusters and mortalities of cardiovascular diseases and extrahepatic cancers, hepatocellular carcinoma and liver related diseases in males and females with SLD.

|  | Extrahepatic<br>cancers |  | Cardiovascular<br>disease |  | HCC |  | Liver related<br>events |  |
| --- | --- | --- | --- | --- | --- | --- | --- | --- |
| Clusters | Events/total | HR (95%CI) | Events/total | HR (95%CI) | Events/total | HR (95%CI) | Events/total | HR (95%CI) |
| Males |  |  |  |  |  |  |  |  |
| < 2 LTCs | 4297 / 93404 | Reference | 2241 / 93404 | Reference | 119 / 93404 | Reference | 439 / 93404 | Reference |
| >=2 LTC | 1595 / 20183 | 1.45 (1.37, 1.54) | 1590 / 20183 | 2.73 (2.56, 2.92) | 56 / 20183 | 1.67 (1.20, 2.32) | 208 / 20183 | 1.84 (1.55, 2.18) |
| Respiratory | 331 / 5587 | **1.24 (1.11, 1.39)** | 254 / 5587 | **1.80 (1.58, 2.05)** | 11 / 5587 | 1.48 (0.80, 2.76) | 45 / 5587 | 1.65 (1.22, 2.25) |
| Mental | 177 / 2814 | **1.30 (1.12, 1.51)** | 159 / 2814 | **2.14 (1.82, 2.52)** | 8 / 2814 | 1.85 (0.89, 3.83) | 39 / 2814 | 2.24 (1.60, 3.14) |
| Cancer | 618 / 6060 | **1.77 (1.62, 1.92)** | 332 / 6060 | **1.83 (1.63, 2.06)** | 14 / 6060 | 1.30 (0.74, 2.27) | 59 / 6060 | 1.69 (1.28, 2.22) |
| Stroke | 163 / 1632 | **1.65 (1.41, 1.94)** | 175 / 1632 | **3.34 (2.86, 3.90)** | 4 / 1632 | 1.30 (0.47, 3.56) | 13 / 1632 | 1.32 (0.76, 2.31) |
| Heart | 306 / 4090 | **1.25 (1.11, 1.41)** | 670 / 4090 | **5.28 (4.83, 5.77)** | 19 / 4090 | **2.44 (1.49, 4.02)** | 52 / 4090 | 2.20 (1.64, 2.96) |
| Females |  |  |  |  |  |  |  |  |
| < 2 LTCs | 1961 / 48930 | Reference | 634 / 48930 | Reference | 17 / 48930 | Reference | 153 / 48930 | Reference |
| >=2 LTC | 966 / 15819 | 1.40 (1.30, 1.52) | 532 / 15819 | 2.30 (2.04, 2.58) | 8 / 15819 | 1.02 (0.43, 2.43) | 77 / 15819 | 1.41 (1.06, 1.86) |
| Respiratory | 228 / 4116 | **1.34 (1.17, 1.54)** | 113 / 4116 | **1.97 (1.61, 2.41)** | 4 / 4116 | 2.19 (0.72, 6.67) | 14 / 4116 | 1.04 (0.60, 1.80) |
| Mental | 175 / 3469 | **1.26 (1.07, 1.47)** | 76 / 3469 | **1.70 (1.34, 2.16)** | 0 / 3469 | NA | 18 / 3469 | 1.54 (0.94, 2.52) |
| Cancer | 265 / 3396 | **1.74 (1.53, 1.98)** | 101 / 3396 | **1.98 (1.60, 2.44)** | 1 / 3396 | 0.61 (0.08, 4.63) | 18 / 3396 | 1.50 (0.92, 2.46) |
| Heart/stroke | 141 / 1745 | **1.65 (1.38, 1.96)** | 168 / 1745 | **5.66 (4.75, 6.75)** | 1 / 1745 | 0.98 (0.12, 7.66) | 13 / 1745 | 2.01 (1.13, 3.57) |
| Thyroid | 157 / 3093 | **1.11 (0.95, 1.31)** | 74 / 3093 | **1.59 (1.25, 2.02)** | 2 / 3093 | 1.26 (0.29, 5.51) | 14 / 3093 | 1.27 (0.73, 2.20) |

Model was stratified by region and age groups, adjusted for ethnicities, education, Townsend Deprivation Index, physical activity, alcohol intake and smoking. LTC: long-term condition. HCC: hepatocellular carcinoma.

## Discussion

In this prospective cohort including 36002 participants with SLD and extrahepatic multimorbidity, we identified five distinct disease clusters for males and females, respectively. Both sexes shared the respiratory cluster, mental health cluster, cancer/osteoarthritic cluster and cardiovascular cluster(s). Males exhibited two separate cardiovascular clusters (i.e., heart cluster and stroke cluster), whereas females had a single combined heart/stroke cluster. A thyroid cluster was only observed in females. Extrahepatic cancers were the leading cause of death across most clusters, except for the cardiovascular clusters, where CVD was the predominant cause. MM was associated with higher overall mortality, with the cardiovascular clusters exhibiting the highest mortality. MM was also associated with increased mortality from extrahepatic cancers, CVD, HCC and liver related diseases, although the strength of these associations varied across clusters.

[comparison with previous studies]

Similar disease clusters have been reported in previous studies, although differences existed due to variations in methods, disease lists, population characteristics and data sources.

Calvin et al. [11], identified 7 clusters in females and 6 in males, including respiratory and cancer clusters in both sexes, and a thyroid cluster uniquely in females, which aligned with our findings. Additionally, their cardiometabolic cluster, characterised with hypertension, coronary heart disease, diabetes and stroke (in males only), closely resembled our cardiovascular clusters. Although we excluded hypertension and diabetes from the list of LTCs due to their inherent link to SLD, we observed a high prevalence of these conditions in the cardiovascular clusters.

Krauth et al. [12] identified five clusters in people aged 40-54 years old and four clusters in people aged 55-70 years old. They found respiratory, mental health, cancer and cardiometabolic clusters, which were similar to our findings. Their cancer cluster was characterised with cancer, arthritis and thyroid diseases, corresponding to our cancer/osteoarthritis cluster and thyroid cluster in females. However, they did not examine sex difference. Both Calvin et al [11] and Krauth et al. [12] used only baseline self-report data in UK Biobank, potentially missing diagnoses captured in hospital records data.

Other UK-based studies reported overlapping clusters. For example, Alessandra Bisquera et al. [20] using multiple correspondence analysis on 32 conditions in UK primary care data, identified 5 clusters, two of which (mental health and cancer/osteoarthritis clusters) were consistent with our findings. Other studies using different clustering techniques also identified relevant patterns. Bodhayan Prasad et al. [21] applied multiple correspondence analysis on 17 diseases in UK Biobank data and identified 4 stroke-centred clusters, while Robertson et al. [22] used clustering analysis on 30 conditions in unselected hospitalised patients in Scotland and has identified 10 clusters.

Outside UK, disease clusters also varied by population. Marina Guisado-Clavero et al. [23] found 5 specific clusters in Spanish population: musculoskeletal, endocrine-metabolic, digestive, neurological, and cardiovascular patterns. A Chilean study [24] identified broad clusters, including a broad depression/CVD/cancer group. A Chinese study [25] manually identified clusters including hypertension, diabetes, coronary heart disease, COPD and stroke, which showed the substantial impact on mortality.

A systematic review [26] highlighted the variability in multimorbidity clusters across studies, but noted common clusters, including mental health and cardiometabolic clusters, both of which we identified. Respiratory cluster was also partially replicable. These comparisons emphasize that while disease clustering differed across studies, certain patterns, particularly cardiometabolic and mental health clusters, consistently emerge.

[cancer/osteoarthritis]

We identified a cancer/osteoarthritis cluster in both males and females, similar to Krauth et al.’s findings [12]. However, the relationship between osteoarthritis and cancers remains inconclusive. Rosas et al. reported osteoarthritis increased prostate cancer progression in animal models [27]. In a Spanish study, Turkiewicz et al. found null associations between osteoarthritis and most cancers, except for a negative association with colorectal cancer [28]. In contrast, Ward et al. observed that osteoarthritis was associated with higher risk of breast, uterus and prostate cancers, and lower risk of digestive tract, lung and ovary cancers in US population [29]. Beydon et al. found that rheumatoid arthritis patients had higher risk of lung, bladder, cervix and prostate cancers, but lower risks of cancers of pancreas, breast and endometrial cancers in French population [30]. Mendelian randomization studies suggested a positive association between osteoarthritis and bladder cancer [31], and ovarian and breast cancers in females [32]. Notably, most prior research has been conducted in the general population, and little is known about this relationship in population with SLD. It is possible that SLD interacts with osteoarthritis, further influencing cancer risk. Further investigations are needed to clarify the direction of these associations and explore potential mechanisms.

[sex differences in clusters]

In this study, 11.0% females were assigned to the heart/stroke cluster, while 20.3% and 8.1% of males to the heart and stroke clusters, respectively, totalling 28.4% of males to the cardiovascular clusters. This aligns with existing evidence showing a higher prevalence and incidence of CVD in males than in females, as reported in the Global Burden of Disease study [33]. This sex gap in CVD is projected to continue widening [34]. Previous studies have emphasized the importance of sex in understanding cardiovascular risk factors [35] and CVD progression [36].

The heart and stroke clusters in males demonstrated distinct characteristics. Compared to the heart cluster, the stroke cluster males were more likely to be younger, more deprived, smoker, physically inactive, hypertension, but less likely to have diabetes and low HDL, as well as a lower mortality rate (26.9 vs. 30.1 per 1000 person-year). Stroke cluster also had higher mortality of cancer, but lower mortality of CVD, compared to the heart cluster.

A mental health cluster was identified in both males and females, characterised primarily by depression and anxiety. However, substance use disorder was present only in the male cluster. This aligns with previous evidence indicating a significantly higher prevalence of substance use disorder in males than in females [37]. Biological, psychological and social factors have been proposed to contribute to these sex differences in substance use and addition patterns women [38].

[risk factors for clusters]

We observed substantial variation in age, socioeconomic status, and lifestyle factors across the derived clusters. Individuals in the mental health and respiratory clusters were general younger than those in cancer or cardiovascular clusters. This likely reflect the earlier onset of mental health and respiratory conditions compared to cancers and CVDs. Both the cancer and cardiovascular clusters were associated with lower educational attainment and greater socioeconomic deprivation. Diabetes and low HDL were also significantly associated with cardiovascular clusters in both males and females. We also observed differences by ethnicity. Asian individuals had lower odds of being in the mental health cluster, whereas Black males also had higher odds of cancer cluster, consistent with previous observations [39–41].

[causes of death and associations with mortality]

We found that MM had higher risk of mortality, consistent with previous evidence [9,42]. In people with SLD, extrahepatic MM increased mortality of CVD, extrahepatic cancers, HCC and liver related diseases. Prior research has highlighted the role of multimorbidity in increasing risk of dementia [11] and cancers [43]. Given that MM also reflects general health status, our findings suggested that overall health status was associated with these mortality outcomes.

In our population with SLD, all disease clusters were linked to elevated mortality, with the highest mortality observed in the cardiovascular-related clusters. Krauth et al. [12] identified 5 disease clusters in UK Biobank cohort, but found increased mortality in only the cardiometabolic and cancer clusters, with the highest mortality in the cancer cluster, which differed from our findings. This discrepancy may suggest that having SLD amplified the adverse effects of multimorbidity, especially cardiovascular clusters. However, other studies have similarly reported high mortality associated with cardiovascular clusters. for example, Swain et al. [44] identified five clusters, and found that cardiovascular-musculoskeletal cluster was associated with the highest mortality, GP consultations and hospitalisation rate, followed by the cardiovascular cluster. A Hong Kong study [45] found that cardiovascular clusters (stroke-hypertension and diabetes-hypertension clusters) were associated with substantially higher mortality rate among eight clusters. Taken together, these findings support that cardiovascular clusters are likely associated with the highest mortality risk.

[implications]

Our findings highlight the heterogeneity of multimorbidity patterns in individuals with SLD and their differential impact on overall and cause-specific mortality. The identification of distinct disease clusters suggests that a one-size-fits-all approach to managing multimorbidity in SLD may be insufficient. Instead, tailored interventions targeting specific clusters may be more effective in improving outcomes, such as cardiovascular risk reduction in the heart and stroke clusters, mental health support in the mental health cluster, and cancer care in the cancer cluster. The particularly high mortality associated with cardiovascular-related clusters underscores the need for early identification and aggressive management of cardiovascular comorbidities in SLD populations. These findings call for further research into the mechanisms driving multimorbidity in SLD and highlight the importance of integrating multimorbidity management into SLD care pathways. Given the observed sex differences in cluster distribution, future strategies should also consider sex-specific approaches to multimorbidity prevention and treatment.

[strengths and limitations]

Our study has several strengths, including a large sample size, long follow-up period, and comprehensive phenotyping of multimorbidity. However, several limitations should be acknowledged. First, we used the FLI to indicate liver steatosis rather than imaging or histological evidence, which may introduce misclassification bias. Second, our multimorbidity phenotyping relied on self-reported data and hospital records, excluding primary care data, potentially leading to an underrepresentation of milder disease cases. Third, UK Biobank participants tend to be predominantly white, less socioeconomically deprived, and healthier than the general population [46], limiting the generalizability of our findings to more diverse or disadvantaged populations. Lastly, we did not account for the temporal sequence of disease development, which may influence the clustering patterns observed.

## Conclusion

In this large cohort study of individuals with SLD and extrahepatic multimorbidity, we identified distinct disease clusters that differed by sex and were associated with varying mortality risks. Cardiovascular-related clusters had the highest mortality, highlighting the need for targeted prevention and management strategies. Our findings underscore the complex interplay between multimorbidity and SLD, emphasizing the importance of a holistic, patient-centred approach to risk stratification and clinical care. Further research is needed to explore the underlying mechanisms and potential interventions to mitigate the burden of multimorbidity in this population.

## Data Availability

All data produced are available online at UK Biobank

## Statements Acknowledgement

We sincerely thank the UK Biobank participants and staff for their contribution to this valuable data resource. This study was conducted under application number 74018.

## Data sharing statement

UK Biobank data are available to registered researchers at https://www.ukbiobank.ac.uk/.

## Author contribution

QF conceived the research idea, conducted data analysis and drafted the manuscript. All authors interpreted results, critically reviewed and revised the manuscript.

